# Charting the Fourth Wave: Geographic, Temporal, Race/Ethnicity, and Demographic Trends in Polysubstance Fentanyl Overdose Deaths in the United States, 2010-2021

**DOI:** 10.1101/2022.11.04.22281945

**Authors:** Joseph Friedman, Chelsea Shover

## Abstract

**Aims:** To characterize polysubstance death in the United States during the transition to the fourth wave of the overdose crisis. To characterize co-involved substances in fatal overdose involving synthetic opioids (mainly illicitly manufactured fentanyl analogs) by year, state, and intersectional sociodemographic groups.

**Design:** Population-based cohort study of national death records.

**Setting:** United States.

**Participants/Cases:** People who died from drug overdose in the United States between 2010-2021.

**Measurements:** Percentage of all fatal overdose involving fentanyls, stimulants, and other drugs. Most commonly co-involved substances in fentanyl overdose by state and year. Percentage of fatal fentanyl overdose co-involving stimulants by state and year. Percentage of fatal fentanyl overdose co-involving stimulants by intersectional region, race/ethnicity, age, and sex.

**Findings:** The percent of US overdose deaths involving both fentanyl and stimulants concurrently rose nearly 60-fold, from 0.6% (n=235) in 2010 to 32.3% (n=34,424) in 2021. In 2010, fentanyl was most commonly found alongside prescription opioids, benzodiazepines, and alcohol. In the Northeast this shifted to heroin-fentanyl co-involvement in the mid-2010s, and nearly universally to cocaine-fentanyl co-involvement by 2021. Universally in the West, and in the majority of states in the South and Midwest, methamphetamine-fentanyl co-involvement predominated by 2021. The proportion of stimulant involvement in fentanyl-involved overdose deaths rose in virtually every state 2015-2021. Cluster analysis reveals particularly high rates for older Black and African American individuals living in the West.

**Conclusions:** By 2021 stimulants were the most common drug class found in fentanyl-involved overdoses in every state in the US. The rise of deaths involving cocaine and methamphetamine must be understood in the context of a drug market dominated by illicit fentanyls, which have made polysubstance use more desirable and commonplace. The widespread concurrent use of fentanyl and stimulants, as well as other polysubstance formulations, presents novel health risks and public health challenges. Ongoing nuanced surveillance is needed to track this rapidly shifting phenomenon.

## Introduction

The United States overdose crisis has escalated in an exponential fashion for over four decades, yet with a shifting profile of drugs implicated in each successive ‘wave’ of the crisis.^1^ Prior to 2007, cocaine was the most commonly involved drug in overdose deaths. This shifted to prescription opioids during 2007-2013 (known as the ‘first wave’ of the overdose crisis), heroin 2014-2014 (“second wave”) and illicit fentanyl analogues 2016-present (“third wave”). Recently, scholars have argued that the ‘fourth wave’ of the US overdose crisis has begun, in recognition of rapidly rising *polysubstance* overdose deaths involving illicitly manufactured fentanyls, with stimulants playing a key role^2–4^. A wide range of polysubstance formulations have been noted in drug checking and overdose mortality data, with myriad substances implicated across numerous drug classes^5–7^. However, the specific details are these shifts remain poorly characterized; more evidence is needed about exact geographic, temporal, race/ethnicity, and demographic trends, as well as which emerging polysubstance formulations are most commonly involved in fatalities. Here we leverage the latest complete data, with records through 2021, to provide a detailed characterization of emerging trends in polysubstance deaths. We focus on fatal polysubstance overdose involving fentanyl and its analogs, as this has become the most common class of drugs involved in fatal overdose in the United States and is understood to be the single most important driver of the current crisis.

## Methods

We obtained data from the Centers for Disease Control and Prevention’s Wide-Ranging Online Database for Epidemiologic Research (WONDER) from 2010 through 2021. Data for 2021 were provisional and as such may represent a slight undercount or overcount of final numbers^5^. We selected all deaths with underlying cause of overdose, using International Classification of Diseases – 10^th^ Edition (ICD-10) codes X40-44, X60-64, or Y10-14. From these, we selected overdose deaths with multiple cause of death code T40.4 (synthetic opioids excluding methadone, a category that is primarily fentanyl and fentanyl analogs). We examined co-involved substances including methamphetamine (T43.6, psychostimulants with abuse potential^8^), cocaine (T40.5), any stimulant (T43.6 or T40.5), benzodiazepines (T42.4), alcohol (T51), prescription opioids (T40.2-3, other opioids), and methadone (T40.3).

We measured the annual percentage of overdose deaths that involved 1) fentanyl, 2) stimulants, 3) fentanyl and stimulants, and 4) neither fentanyl nor stimulants. We then measured the most commonly co-involved substance at the state level over the study period. For intersectional analysis, in order to have a large enough sample to simultaneously stratify by census region, race/ethnicity, sex, and age group, we combined all fentanyl deaths over the most recent five-year period with finalized data, 2016-2020.

## Results

The polysubstance characteristics of fentanyl-involved overdose mortality shifted dramatically throughout the 2010 to 2021 period. As overdose deaths rose in the United States from 38,643 in 2010 to 106,719 in 2021, the percent involving both fentanyl and stimulants concurrently rose nearly 60-fold, from 0.6% (n=235) to 32.3% (n=34,424) (**Figure 1**). The proportion of deaths involving fentanyl without stimulants also rose from 7.2% in 2010 to a peak of 35.7% in 2020, before declining slightly to 33.9% in 2021. The proportion with stimulants and no fentanyl remained relatively more stable, from 14.8% in 2010 to 17.9% in 2021. The proportion containing neither fentanyl nor stimulants fell from 77.3% in 2010 to only 16.0% in 2021.

**Figure 1.**
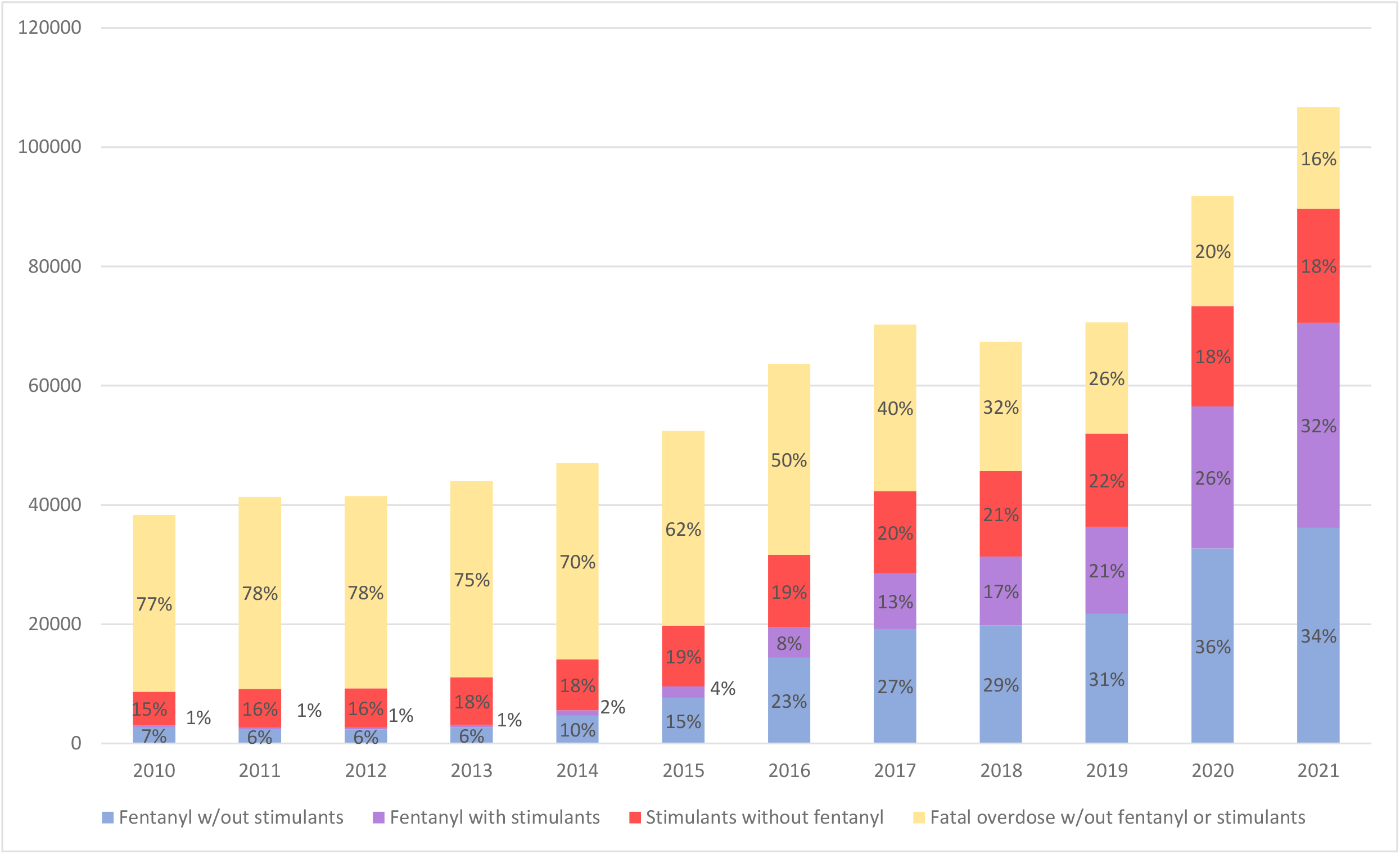
Overdose Deaths by Fentanyl and Stimulant Presence, 2010-2021.

In 2010, fentanyl was most often co-involved with prescription opioids (19 states), alcohol (18 states) and benzodiazepines (eight states), with that general pattern seen across all four major census regions (**Figure 2**). This pattern shifted earliest in the Northeast, as heroin became the most common co-involved substance in 2014 in five states (of nine total in the region). Cocaine became the most commonly co-involved substance among states in the Northeast in 2019 with seven states. By 2021, all states in the Northeast had a stimulant as the most common co-involved substance, with seven having cocaine and two having methamphetamine. Among states in the West region, a mixture of prescription opioids, benzodiazepines, and alcohol were the most common co-involved substances through 2020, when methamphetamine was the most common in ten states (of 13 in the region), which grew to all 13 states by 2021. The Midwest and the South saw a more mixed profile, with brief periods of heroin/fentanyl predominance in numerous states between 2016 and 2018. By 2021, methamphetamine and cocaine were the only leading co-involved substances represented in these regions, with 19 and 10 respectively of a total 28 states and the District of Columbia.

**Figure 2.**
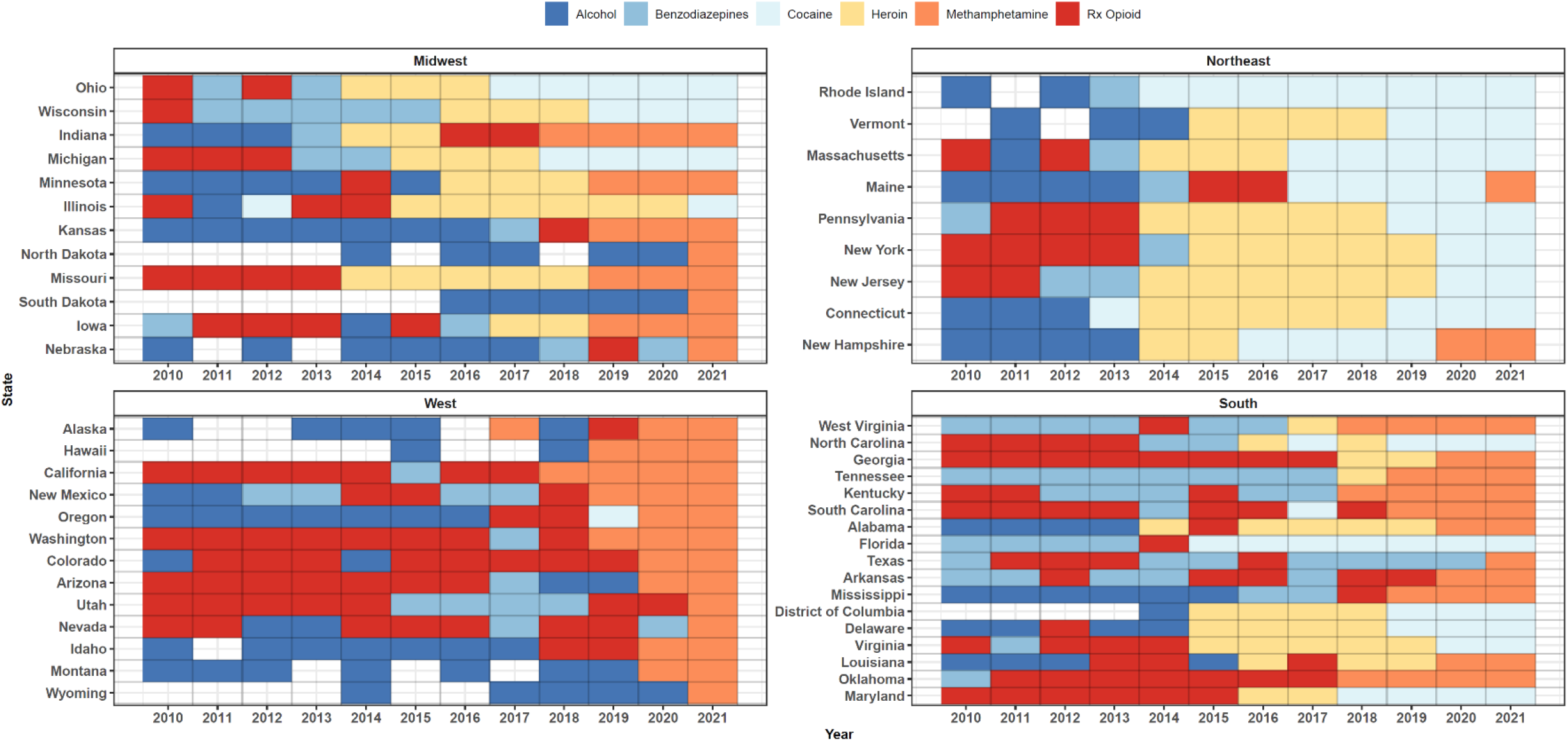
Most Common Drug Co-Involved in Overdose Mortality With Fentanyls, by State and Year, 2010-2021.

In 2021, methamphetamine co-involvement was highest in states in the Pacific (California, Oregon, Washington, Idaho, and New Mexico), as well as West Virginia and Kentucky (**Figure 3)**. Cocaine co-involvement was highest in Rhode Island, Vermont, and Massachusetts, as well as a broad swath of states in the Northeast and Southeast. Benzodiazepine co-involvement in 2021 was highest in Texas (30.2%),Utah (28.8%), Arkansas (22.1%), and Massachusetts (22.0%).

**Figure 3.**
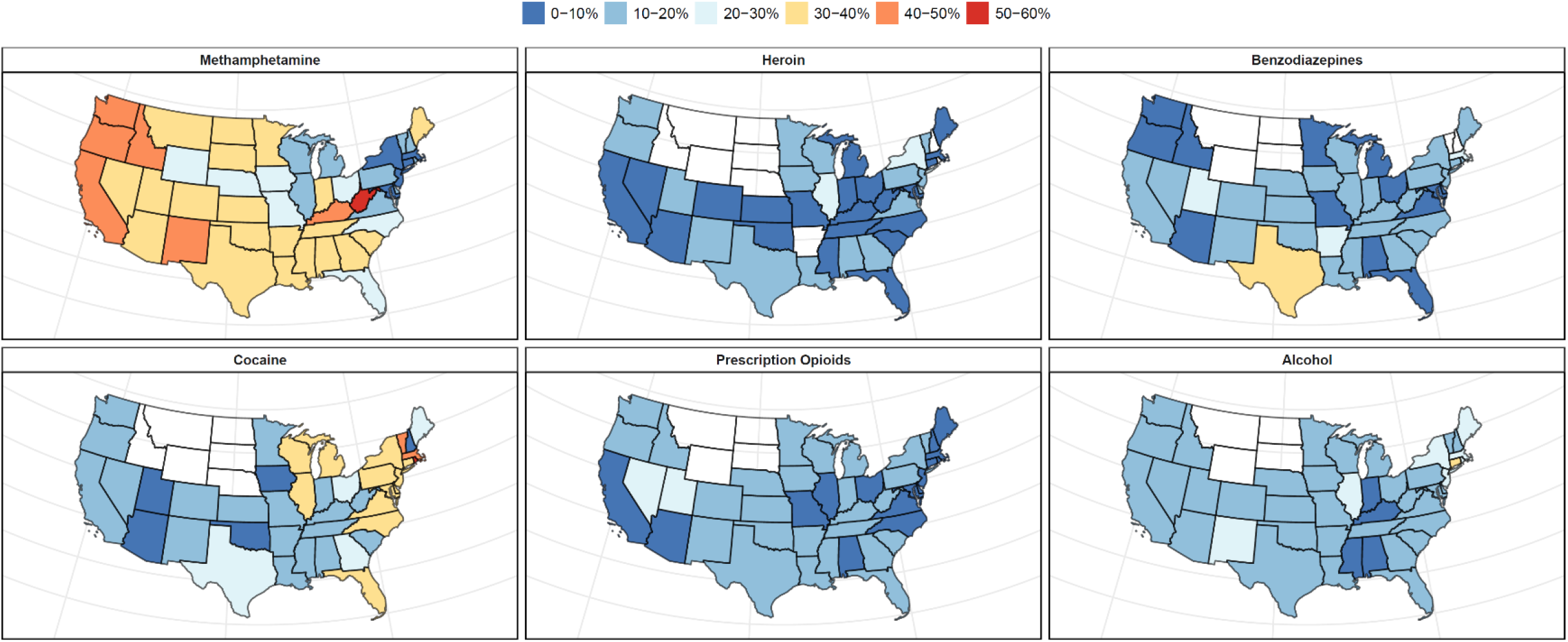
Percent of Fentanyl Overdose Deaths Containing Other Drug Classes by State, 2021.

Virtually all states observed an increase in the proportion of fentanyl deaths involving stimulants between 2015 and 2021 (**Figure 4**). By 2021, the states with the highest proportions included Alaska (65.1%), Hawaii (60.4%), West Virginia (60.2%), Rhode Island (58.8%) and California (58.5.X%) (**Figure 4**). The states with the lowest proportions in 2021 included New Hampshire (19.2%), Nebraska (30.4%), and Wyoming (30.6%).

**Figure 4.**
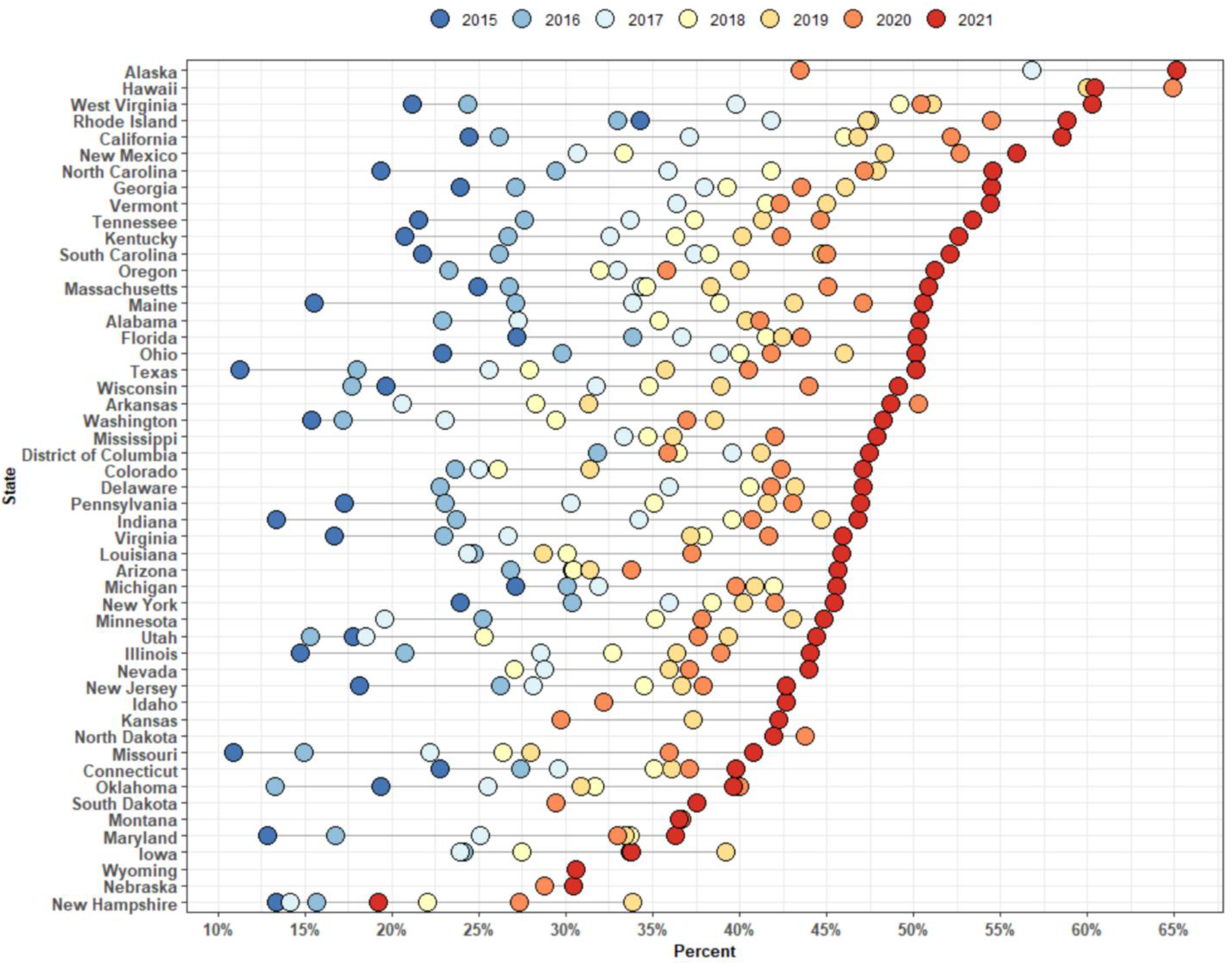
Percent of Fentanyl Overdose Deaths Involving Stimulants by State and Year, 2015-2021.

**Figure 5** shows the proportion of fentanyl deaths containing stimulants for clusters defined by the intersection of census region, race/ethnicity, gender, and 10-year age groups, between 2017 and 2021. **Table 1** also highlights trends separate by census division, age, gender, census division, race, and ethnicity in 2021. Both overall, and in specific clusters, the highest prevalence of stimulant involvement in fentanyl overdose deaths was observed in individuals aged 25 through 59, with generally lower rates among the youngest and oldest individuals. The clusters with the highest proportions included 65–74-year-old Non-Hispanic Black or African American women living in West (73.3%), as well as 55–65-year-old Black or African American Men living in the West (68.7%).

**Table 1.** Percent of Fentanyl Overdose Deaths Containing Stimulants by Key Sociodemographic Characteristics, 2021.

|  | Fentanyl | Fentanyl and<br>stimulants | % Fentanyl deaths with<br>stimulants |
| --- | --- | --- | --- |
| **Sex** |  |  |  |
| Male | 51022 | 24628 | 48.3% |
| Female | 19567 | 9796 | 50.1% |
| **Race/Ethnicity** |  |  |  |
| Non-Hispanic |  |  |  |
| American Indian or Alaska Native | 804 | 421 | 52.4% |
| Asian | 464 | 228 | 49.1% |
| Black or African American | 13590 | 7233 | 53.2% |
| Native Hawaiian or Pacific Islander | 57 | 27 | 47.4% |
| More than one race/ethnicity | 815 | 388 | 47.6% |
| White | 45587 | 21924 | 48.1% |
| Hispanic |  |  |  |
| American Indian or Alaska Native | 86 | 46 | 53.5% |
| Asian | 26 | 0 | 0.0% |
| Black or African American | 316 | 151 | 47.8% |
| Native Hawaiian or Pacific Islander | 19 | 11 | 57.9% |
| More than one race/ethnicity | 149 | 71 | 47.7% |
| White | 8252 | 3914 | 47.4% |
| **Age group** |  |  |  |
| <14 years | 140 | suppr. | -- |
| 14-18 | 849 | 135 | 15.9% |
| 19-24 | 5110 | 1803 | 35.3% |
| 25-29 | 8075 | 3579 | 44.3% |
| 30-39 | 20713 | 10435 | 50.4% |
| 40-49 | 15500 | 8416 | 54.3% |
| 50-59 | 13121 | 7002 | 53.4% |
| 60-69 | 6335 | 2803 | 44.2% |
| 70-79 | 654 | 197 | 30.1% |
| >79 | 13 | suppr. | -- |
| **Census Division** |  |  |  |
| New England | 4840 | 2242 | 46.3% |
| Middle Atlantic | 10762 | 4878 | 45.3% |
| East North Central | 12483 | 5911 | 47.4% |
| West North Central | 3085 | 1274 | 41.3% |
| South Atlantic | 17690 | 8789 | 49.7% |
| East South Central | 5788 | 3027 | 52.3% |
| West South Central | 3592 | 1713 | 47.7% |
| Mountain | 4163 | 1952 | 46.9% |
| Pacific | 8186 | 4638 | 56.7% |

**Figure 5.**
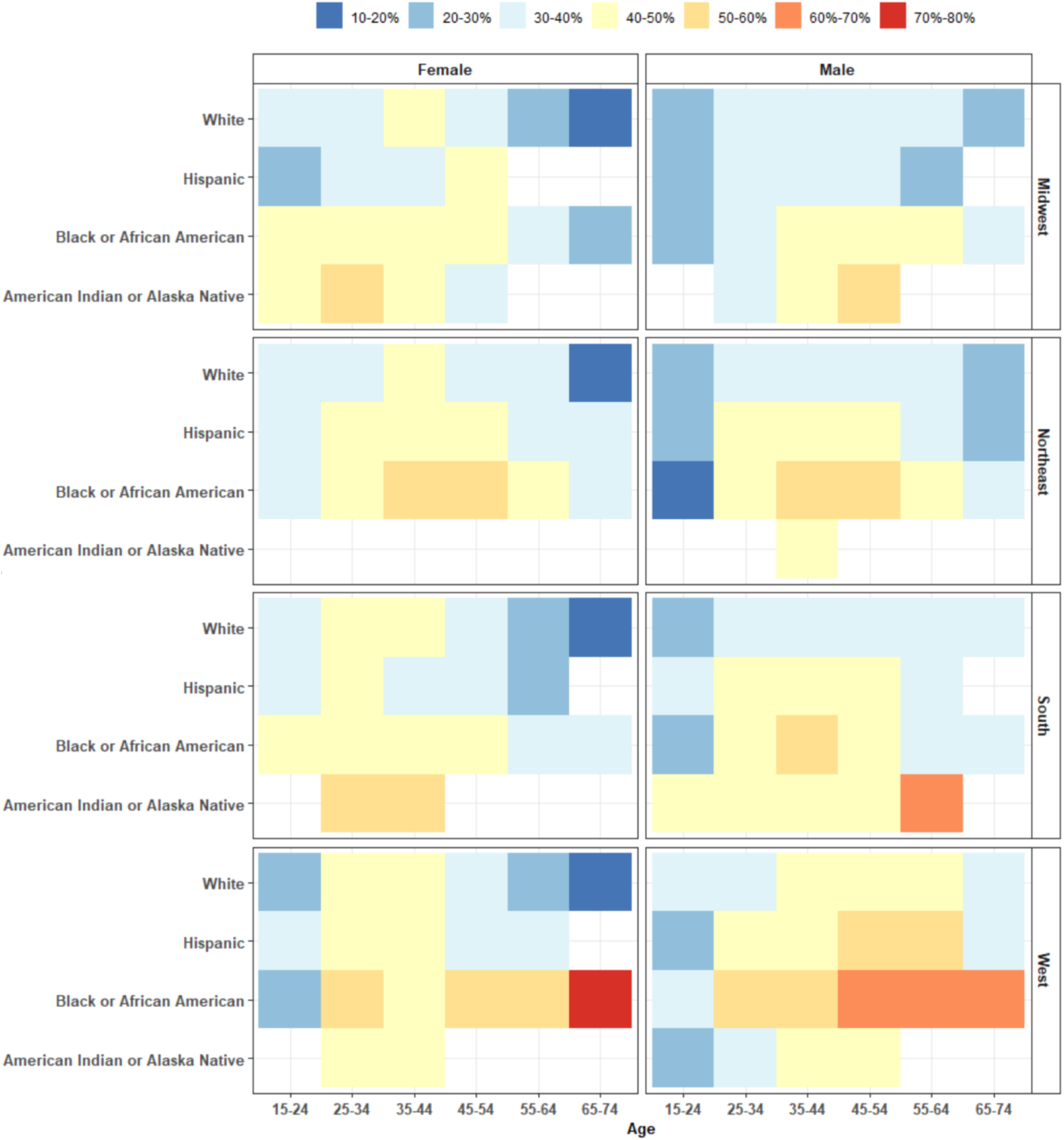
Percent of Fentanyl Overdose Deaths Involving Stimulants by Intersectional Region, Race/Ethnicity, Age, Gender, 2016-2020.

Although men represented a much larger share of all fentanyl overdose deaths, with 51,022 among men vs. 19,567 among women in 2021, they had largely comparable rates of stimulant co-involvement (48.3% among men and 50.1% among women). Among census divisions, the proportion of fentanyl deaths involving a stimulant ranged from 41.3% in the West North Central division, to 52.3% in the East South-Central division.

## Discussion

The rise of illicitly manufactured fentanyls has ushered in an overdose crisis in the United States of unprecedented magnitude^2,9–11^. This has created conditions that promote a number of other shifts in the illicit drug supply, leading to rising polysubstance overdose deaths—the so-called ‘fourth wave’ of the crisis^2,5^. Mixtures of fentanyl analogues and drugs of various drug classes, such as stimulants, benzodiazepines, tranquilizers, and other opioids have been noted in distinct geographies^2,5,6,12^. Here we provide a comprehensive characterization of the rising fentanyl-based polysubstance overdose crisis, detailing shifts in specific drug combinations over time, and which geographic, racial/ethnic, and demographic groups are the most affected.

In 2010, fentanyl was most commonly found alongside prescription medication (opioids and benzodiazepines) and alcohol, i.e. largely products produced in legal markets. Over the past decade this has shifted first to heroin-fentanyl combinations in specific states, and then universally to illicit stimulants. The fraction of all overdose deaths involving both fentanyl and stimulants grew 60-fold between 2010 and 2021, and is on track to represent the single largest component of the overdose crisis in the near future. However, this has occurred in a distinct fashion based on geography and time. The northeastern states nearly universally saw a distinct period of heroin-fentanyl co-involvement, which was also found in some parts of the Midwest and South but was completely absent from the Western states (which transitioned rapidly from black tar heroin to fentanyl with methamphetamine co-involvement). By 2021, cocaine predominated in the Northeast and methamphetamine had become the most common drug found alongside fentanyls in the rest of the country.

There is also a considerable variation between regions in the total degree of stimulant co-involvement, including a nearly three-fold difference seen between states in 2021. The most affected states have been in the Pacific (with methamphetamine involvement) and in the Northeast (with cocaine involvement). Cluster analysis reveals particularly high rates for older Black and African American individuals living in the West.

The rise of deaths involving cocaine and methamphetamine must be understood in the context of a shifting illicit opioid drug market increasingly dominated by illicit fentanyls^10^. Recent ethnographic and qualitative research suggests that fentanyls have created conditions that make polysubstance use more desirable and commonplace^13,14^. For instance, many individuals report that mixing a small amount of methamphetamine into injected doses of fentanyl subjectively prolongs the onset of withdrawal symptoms, increases euphoria, decreases overdose risk, and improves energy levels required to continue to collect funds for the next set of drug purchases^13–15^. These perceived advantages may be particularly important given the short duration of fentanyls, requiring individuals to inject far more frequently than heroin, and the heightened overdose risk from each injection^10^. Similar findings have been reported in qualitative studies of xylazine and other drugs commonly added to fentanyls, suggesting a possible structural similarities across various emerging polysubstance patterns^5^. Given the increased risk of negative health outcomes such as naloxone-resistant overdose, precise surveillance of specific drug formulations and sociodemographic groups affected is essential^5^.

### Limitations

There are limitations to this study that should be considered. As we highlight here, the landscape of polysubstance overdose has been evolving in a highly rapid manner. Therefore, even the most current results may simply represent snapshots of shifting dynamics that will soon change. Further, fatal overdose is the most readily available metric, as there are highly limited toxicological surveillance data on non-fatal overdose in the United States. However, they don’t represent the totality of use practices, rather the fraction of drug use most likely to result in death. The categories of drugs assessed here are also limited by current CDC classification schemes, which limit within-category assessment. For instance, is it not possible to distinguish between prescription benzodiazepines and novel synthetic benzodiazepines, or between fentanyl analogues and nitazines. Finally, particularly for the cluster analysis, stratification has led to small sample sizes for some analytical units, although samples sizes for most of the analysis were robust.

### Conclusions

We provide a detailed description of the fourth wave of the US overdose crisis—characterized by sharply rising polysubstance overdose deaths involving illicitly manufactured fentanyls. Stimulant-fentanyl co-involvement is rapidly becoming the largest component of the crisis, with a distinct pattern seen by over time and by geography and sociodemographic groups. The regional patterning of cocaine-fentanyl in the Northeast and methamphetamine-fentanyl in the rest of the country is particularly notable. The widespread concurrent use of fentanyl and stimulants, as well as other polysubstance formulations, presents numerous novel health risks and public health challenges. Moving forward, ongoing nuanced surveillance is needed to track this rapidly shifting phenomenon.

## Data Availability

All data produced in the present study are publicly available online at: https://wonder.cdc.gov/mcd-icd10.html and https://wonder.cdc.gov/mcd-icd10-provisional.html. The authors may be contacted for assistance.

https://wonder.cdc.gov/mcd-icd10-provisional.html.

